# The impact of unplanned school closure on children’s social contact: Rapid evidence review

**DOI:** 10.1101/2020.03.17.20037457

**Authors:** Samantha K. Brooks, Louise E. Smith, Rebecca K. Webster, Dale Weston, Lisa Woodland, Ian Hall, G. James Rubin

## Abstract

**Background:** Emergency school closures are often used as public health interventions during infectious disease outbreaks in an attempt to minimise the spread of infection. However, if children continue to mix with others outside the home during the closures, these measures are unlikely to be effective.

**Objectives:** This review aimed to summarise existing literature on children’s activities and contacts made outside the home during unplanned school closures.

**Methods:** We searched four databases from inception to February 2020 for relevant literature. Main findings were extracted.

**Results:** 3,343 citations were screened and 19 included in the review. Activities and social contacts appeared to decrease during closures but contact was still common. All studies reported children leaving the house or being looked after by non-household members. There was some evidence that older child age and parental disagreement with closure were predictive of children leaving the house, and mixed evidence regarding the relationship between infection status and leaving the home. Parental agreement with closure was generally high, but some parents disagreed due to perceived low risk of infection and practical issues regarding childcare and financial impact.

**Conclusions:** Evidence suggests that many children continue to leave the house and mix with others during school closures despite public health recommendations to avoid social contact. This review of behaviour during unplanned school closures could be used to improve infectious disease modelling.

## Introduction

Gaining control of an infectious disease outbreak can require difficult decisions to be made. Children are often in close physical proximity at school, have less than perfect hygiene behaviours and have low prior immunity to many infections, leading them to be dubbed ‘super-spreaders’ of influenza [1]. For this reason, school closures are often mooted as one way of delaying the spread of infection [2]. There is evidence to suggest that social contacts should reduce when schools are closed. For example, it has been reported that students have contact with fewer people at weekends [3] and that the number of contacts children have with others approximately halves during the holidays [4, 5]. Several studies have also examined illness transmission rates during planned school closures, reporting a reduction in illness during school holidays [6, 7, 8] and teacher strikes [9].

School closure is not a step that can be taken lightly, however. Clearly, closures can have an impact on the education of the children involved. But they can also have an impact on the healthcare system and wider economy if large numbers of the workforce stay home to look after their children, on household incomes, on social policies implemented at school and on the likelihood of children engaging in other risky behaviours if they must be left unattended at home [10]. Indeed, the secondary economic and social effects of school closures are potentially huge [11].

Understanding whether the efficacy of school closure in terms of reducing the spread of disease is sufficient to justify these impacts is therefore important. One of the key unknowns that prevents us from resolving this issue is what happens to children after a school is closed. The optimum answer, from an epidemiological perspective, is that they remain in their bedrooms for the duration of the closure, never coming into contact with another person [12, 13]. However, this is impractical and front-line experience of outbreak management is full of tales of children continuing to congregate after being sent home from school and sometimes engaging in behaviour likely to increase the risks of infection spreading [14, 15]. Any full assessment of the impact of school closures must take this into account.

A related issue is the extent to which the quality rather than quantity of social interaction changes following a school closure. For example, the role of grandparents in caring for children following school closure may be a cause for concern if this increases the likelihood of older adults coming into contact with the infectious disease in question.

Finally, given that school closures are often accompanied by advice to parents to limit the contact their children have with others, understanding what practical or attitudinal factors affect the likelihood of children mixing during a closure may also be helpful in improving the advice that is given out.

In this rapid evidence review, we examined a) what is currently known about the impact of unplanned school closure on children’s interaction with others outside the home, b) who provides childcare during a closure, c) what factors are associated with children interacting with others outside the home during a closure, and d) what affected parents think about closures.

## Method

This work was carried out as a rapid evidence review in response to the COVID-19 outbreak in December 2019, which has led to policy-makers worldwide discussing how best to minimise the spread of the disease. As there are no specific guidelines for rapid reviews, the PRISMA checklist has not been completed.

### Search strategy and selection criteria

We used the following search strategy to search MEDLINE®, PsycINFO® and EMBASE®:

1. school* ADJ3 close* OR ADJ3 closure* OR ADJ3 closing* OR ADJ3 dismiss*
2. nurser* ADJ3 close* OR ADJ3 closure* OR ADJ3 closing* OR ADJ3 dismiss*
3. kindergar* ADJ3 close* OR ADJ3 closure* OR ADJ3 closing* OR ADJ3 dismiss*
4. playgroup* ADJ3 close* OR ADJ3 closure* OR ADJ3 closing* OR ADJ3 dismiss*
5. play-group* ADJ3 close* OR ADJ3 closure* OR ADJ3 closing* OR ADJ3 dismiss*
6. 1 OR 2 OR 3 OR 4 OR 5
7. behaviour* OR behavior* OR contact* OR mix* OR social* OR targeted layered containment
8. 6 AND 7

We repeated the same search on Web of Science using NEAR instead of ADJ3.

To be included in the review, studies had to: i) report on primary research; ii) be published in peer-reviewed journals; iii) be written in English or Italian (languages spoken by our team); and iv) report on social activities of children during unplanned temporary school closures (because mixing behaviour will likely be different during closures with plenty of notice, when parents have more time to plan what to do). We excluded papers based on intentions, hypothetical scenarios or simulations.

### Screening

One author (SKB) ran the search strategy on all databases and downloaded all resulting citations to EndNote© version X9 (Thomson Reuters, New York, USA). All titles and abstracts were screened for relevance according to the inclusion criteria by at least two authors (SKB, LES, RKW, DW, or LW). The authors compared which texts they had chosen for inclusion and discrepancies were resolved through discussion with the wider team. Full texts of all remaining citations were obtained and reviewed by one author (SKB), excluding any which did not meet all inclusion criteria. Finally, the reference lists of remaining papers were hand-searched for any additional relevant studies. A flow chart of the screening process is presented in Figure 1.

**Figure 1.**
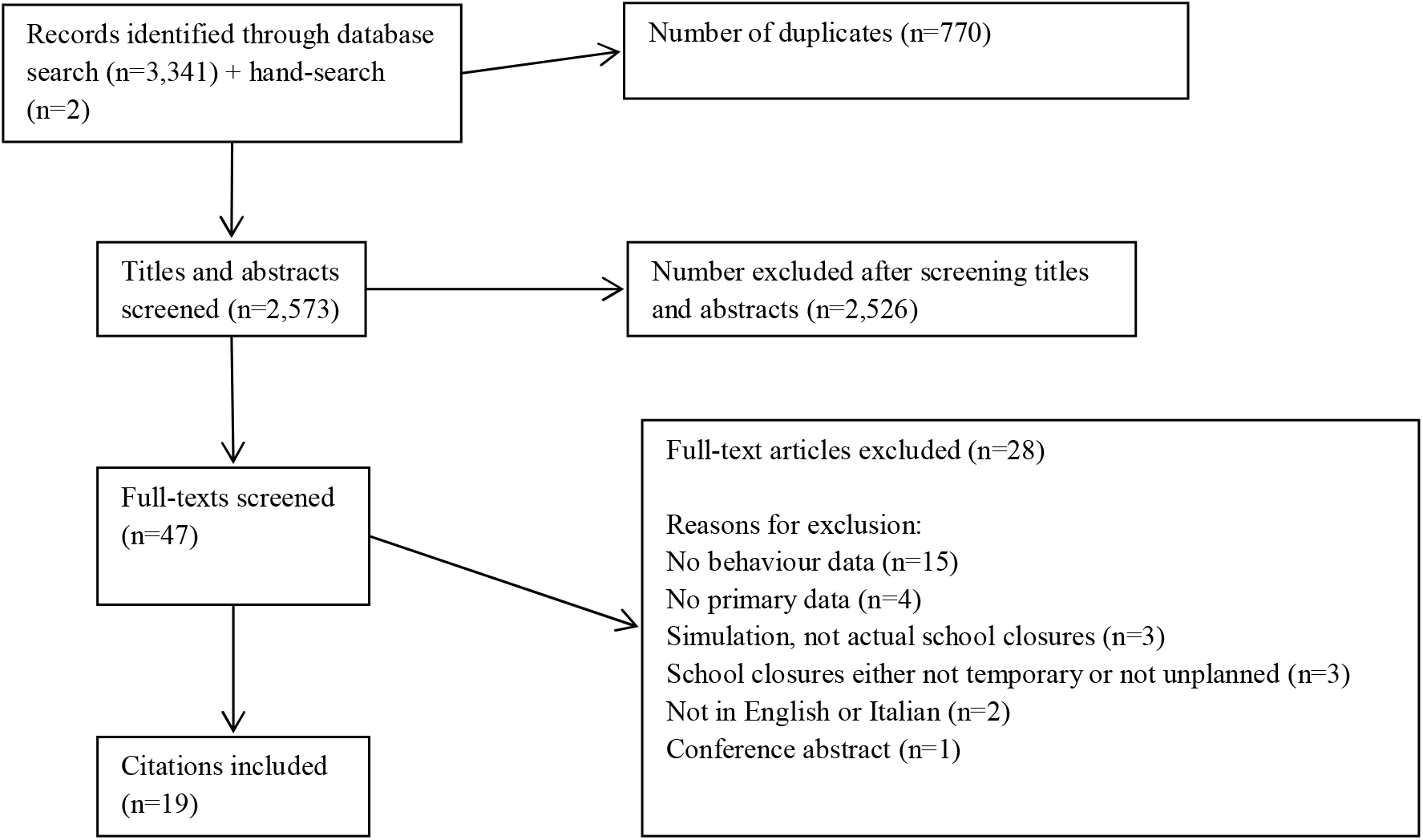
Screening process.

### Data extraction

We designed spreadsheets to extract the following data from papers: authors, publication year, country of study, design, participants (including *n* and demographic information), reason for school closure, length of school closure, and key results. Data extraction was carried out by one author (SKB).

## Results

Nineteen papers were included, eighteen of which used a cross-sectional design using questionnaires to assess difficulties during the school closures, activities outside the home during the closures and/or who provided childcare during the closures. The remaining paper used a qualitative design. The majority (n=10) were from the USA; four papers were from Australia, and the remaining papers from Argentina, Japan, Russia, Taiwan and the UK (one for each country). Most papers reported on school closures due to the H1N1 pandemic (n=11) or other influenza or influenza-like outbreaks (n=7); one paper reported on a school closed in preparation for a hurricane. The duration of school closures ranged from one day to two weeks. The size of the quantitative studies ranged from 35 households (representing 67 children) to 2,229 households (representing 4,171 children). Table 1 provides a summary of activities outside the home found in the reviewed studies, while Table 2 summarises childcare arrangements involving non-household members during school closures. Supplementary Appendix 1 provides more detail on the results of each study.

**Table 1.** Activities outside the home during school closures

| **Reference, country** | **Participants** | **Activities outside the home** |
| --- | --- | --- |
| Basurto-Davila et al. (2013), Argentina [27] | 226 households; Children aged 6-15 from 3 schools closed for 2 weeks due to H1N1 | 67% of children visited public places at least once; 45% left the house several times. |
| Borse et al. (2011), USA [24] | 554 households; Median age of children: 8 years; Schools closed for 5-7 days due to H1N1 | 30% of students visited at least one locale outside their homes. |
| Braunack-Mayer et al. (2013), Australia [14] | 4 school principals, 25 staff, 14 parents, 13 students (age 12-17); Schools either partially or fully closed due to H1N1 (length of closure unclear) | Qualitative study indicating most people adhered to advised quarantine, but in the absence of clear instructions, many invented their own rules.<br>Some parents quarantined their children to avoid being seen as irresponsible. However, many were home alone and so it was unclear whether children complied. Others reported seeing the closure as ineffective and did not quarantine their children. One student reported meeting friends regularly even though his parents believed he was at home. |
| Chen et al. (2011), Taiwan [30] | 232 households; School for children aged 5-12; School closed for 7 days due to H1N1 | 13% went to public places or gatherings at least once,<br>12% visited relatives,<br>5% went to parents' workplace. |
| Effler et al. (2010), Australia [19] | 233 households; Median age of children 11 years (range 5-13); 3 schools closed due to H1N1; School A closed entirely 'for the coming week'; Schools B and C cancelled classes for grades 5 and 5-7 respectively | 74% participated in activities outside the home on at least one occasion, reporting a total of 860 out-of-home activities with an average of 3.7 out-of-home activities for each student. |
| Epson et al. (2015), USA [20] | 35 households, representing 67 students; 1 elementary school and 1 junior and senior high school housed in the same building complex; Schools closed between 29.01.13 and 05.02.13 due to | 58% visited at least one outside venue. |
|  | influenza-like illness |  |
| Gift et al. (2010), USA [23] | 214 households, with 269 children under 18; Elementary school closed for 1 week due to H1N1 | 69% visited at least one other location. |
| Jackson et al. (2011), UK [16] | 107 students (only 46 reported how many times they visited public places during closures); Children aged 11-15; School closed for 1 week, reopened for 2 days, then closed for another week, due to H1N1 | 98% visited more than one place. 73 students provided their typical number of contacts per day during closure and 35 also provided information for a typical school day. Mean totals of reported contacts were 70.3 and 24.8 during typical school days and closure respectively. |
| Johnson et al. (2008), USA [22] | 220 households, with 355 children; Median age of children: 12 (range 5-19); Schools closed for 12 days due to influenza virus B | 89% visited at least one public location and 47% travelled outside of the county. |
| Litvinova et al. (2019), Russia [17] | 450 participants including students and their household members; School A for children aged 6-17 and School B for children aged 6-15; Schools closed for 7 days to mitigate spread of seasonal influenza | There was a reduction in the number of contacts made by students (14.2 contacts per day when open vs. 6.5 when closed). Students significantly reduce their number of contacts with individuals aged under 18 (75% reduction) and 19-59 year-olds (20% reduction), while increasing contacts with individuals aged 60 or over (52% increase), although the absolute value remained low (less than one contact per day). |
| McVernon et al. (2011), Australia [28] | 314 households; 33 schools; Schools with confirmed cases of H1N1 in multiple classes were entirely closed for 7 days while schools with confirmed cases in only one class were instructed to close only that class | 43 households reported that a child spent at least one day outside the family home and mixing with other children occurred on almost half of these occasions (48.8%). Contact with children who were not immediate family members was less likely during days spent at home. No child visited a household in which another child was ill, compared with reported child visitors in 15.9% of 226 homes without a case. |
| Miller et al. (2010), USA [18] | 63 parents of 176 lower school students (grades 5-8); 188 upper school students (grades 9-12); Week-long closure due to H1N1 | Upper school: Mean number of days spent on activities: 3.42 any other outdoor activity; 2.44 eating at restaurants; 1.89 using public transport; 1.48 hosting a friend; 1.47 shopping; 1.47 any other indoor |
|  |  | activity; 0.44 working at a job. Average number of friends seen per day: 2.53 Wednesday, 2.06 Thursday, 2.59 Friday, 2.40 Saturday, 1.23 Sunday, 1.02 Monday, 1.05 Tuesday.<br>Lower school: Mean number of days spent on activities: 2.77 any other outdoor activity; 1.34 eating at restaurants; 1.12 any other indoor activity; 1.05 shopping; 0.73 visiting a friend; 0.55 hosting a friend; 0.10 using public transport. Average number of friends seen per day : 0.30 Wednesday, 0.52 Thursday, 0.84 Friday, 0.83 Saturday, 1.17 Sunday, 0.74 Monday, 0.68 Tuesday. |
| Mizumoto et al. (2013), Japan [25] | 882 households; 25.2% in kindergarten, 24.8% in primary school, 25.1% in junior high school and 24.9% in high school, age range 4-18; 'School closure or class suspension at least once' due to H1N1 | 20.5% left the home for non-essential reasons. |
| Russell et al. (2016), USA [26] | 99 households, representing 197 children; Students in pre-kindergarten up to 12<sup>th</sup> grade; School closed for 4 days due to influenza-like illness | 77% of children went outside the home or visited with a non-household member, participating in a mean of two activities (IQR 1-4). |
| Steelfisher et al. (2010), USA [31] | 523 parents; Ages not reported; Childcare centres and schools closed due to H1N1 - 10% were closed for 1 day, 19% for 2 days, 29% for 3 days, 15% for 4 days, 17% for 5 days, 9% for more than 5, 2% didn't know | 56% reported their child participated in at least one activity involving people outside the household. |
| Timperio et al. (2009), USA [29] | 262 households representing 480 children; Ages not reported; Two schools closed due to seasonal influenza; one closed for 3 days and the other closed for 4 days | 43.3% visited strip malls or Wal-Mart (the largest store in the area); 42.9% visited family; 38.7% went grocery shopping; 32.6% ate at restaurants; 30.3% either visited friends' houses or had friends visiting their house; 29.1% attended religious services; 23.8% took part in sports |
|  |  | activities; 17.6% went to public gatherings such as concerts, movies or festivals; 8.4% went to a part time job. |
| Tsai et al. (2017), USA [32] | 208 households with 423 children; School closed for 8 days due to influenza | Not reported. |
| van Gemert et al. (2018), Australia [21] | 99 students with laboratory confirmed pH1N1; Age 6-17; Seven schools closed for 3-9 days (not including weekends) | 26% (21/81) who reported usually taking part in extra-curricular activities (not sports or religious activities) continued to take part in extra-curricular activities. |
| Zheteyeva et al. (2017), USA [33] | 2,229 households with 4,247 students; Kindergarten – Grade 12; Schools closed for 4 days in preparation for Hurricane Isaac | Not reported. |

**Table 2.** Childcare arrangements involving non-household members during school closures

| **Reference, country** | **Childcare arrangements involving non-household members** |
| --- | --- |
| Basurto-Davila et al. (2013), Argentina [27] | Left with a relative or family friend (82% / 88% depending on region), hired nanny (13% / 5%), other special arrangements (3% / 4%), left alone (2% / 1%). |
| Chen et al. (2011), Taiwan [30] | Parents (60%), other relatives (35%), others (4%), left alone (1%). |
| Effler et al. (2010), Australia [19] | Asymptomatic students: with children other than their siblings (19%). Ill students: with children other than their siblings (6%). All students: left alone for at least some time (10%) |
| Epson et al. (2015), USA [20] | Adult from outside the household (9%), work with parents (6%), childcare program (3%), left alone (9%). |
| Gift et al. (2010), USA [23] | Home as main location (77%). The next most common locations were another family member's home, non-family member's home, parents' workplace, vacation, daycare, 'other'. |
| Jackson et al. (2011), UK [16] | Among caregivers for whom information was available, 125/182 (69%) would have seen the student on a typical school day. |
| Johnson et al. (2008), USA [22] | Special childcare arrangements including grandparents, other relatives, other adults, taking the child to work, having older siblings watch them or using childcare programs (10%), one or more night spent outside the household (3%). |
| McVernon et al. (2011), Australia [28] | Households with influenza: Adult from outside the home (44.4% for households that complied with advice to remain in home vs 2.4% for noncompliant households). Households without influenza: Adult from |
|  | outside the home (28.3% for households that complied vs 4.0% for noncompliant households). |
| Miller et al. (2010), USA [18] | Proportion of caregivers (upper school): 0.62 parent, 0.24 sibling, 0.07 grandparent, 0.07 other, 0.06 nanny / babysitter, 0.07 friend's caretaker, 0.11 other, 0.88 self. Proportion of caregivers (lower school): 0.85 parent, 0.30 sibling, 0.09 grandparent, 0.15 other family, 0.27 nanny / babysitter, 0.03 friend's caretaker, 0.06 other, 0.76 self. |
| Mizumoto et al. (2013), Japan [25] | Another household member (64.3%), left alone (28.5%), special arrangement such as parental absence from work (7.3%). |
| Russell et al. (2016), USA [26] | Adult from outside the household (20%); childcare programme (1%) |
| Steelfisher et al. (2010), USA [31] | 81% were cared for by an adult in the household, 20% by a family member outside the household, 1% by a friend/neighbour, 3% by a professional care provider, and 10% stayed home alone. |
| Tsai et al. (2017), USA [32] | Childcare programme (3%), attending work with parents (1%), left alone without supervision (1%), old enough to care for themselves (15%). |
| Zheteyeva et al. (2017), USA [33] | Old enough to care for themselves (11.6%), went to work with parents (5.3%), childcare program (2.6%), left alone without supervision (2.5%). |

### Interaction with others outside the home

Participation in activities and interactions with others did appear to decrease during school closures compared to regular school days [16, 17, 18]; for example, one study [16] reported that school closure was associated with a 65% reduction in the mean total number of contacts for each student. However, social contact was still common. All nineteen studies showed that at least some (and in some cases, most) children took part in activities outside of the home during school closures, even despite health recommendations to remain indoors and isolated from others.

### Factors associated with contact outside the home

#### Infection status

Several studies suggested that children who reported illness during a school closure were less likely to take part in activities outside the home [16, 19, 20, 21]. For example, Effler et al. [19] reported a significant difference for the proportion of cases (students testing positive for H1N1 virus), students who had been in close physical proximity to cases, and peers who did not meet case or contact criteria who reported leaving the home more than once during the closure period (42%, 66% and 92% respectively). Cases reported an average of 0.8 out-of-home activities per student per week, compared with 2.9 for contacts and 5.6 for peers. Other studies reported that children who reported illness or lived in households in which influenza-like illness was reported did not participate in the majority of activities reported by other students [20, 21] and that their contact with others was reduced [16].

However, other studies reported few differences in out-of-home activities between symptomatic and asymptomatic children [18, 22, 23, 24, 25]. In fact, one study [18] found that students with illnesses were more likely to report an increase in travel plans, for reasons that are not clear. Two studies found that children with an influenza-like illness were more likely to have visited a healthcare provider, but no other statistically significant differences in out-of-home activities were found between students with and without symptoms [23, 24].

#### Age

Three studies noted more activities and contacts among older children [18, 22, 26]. In particular, Grade 12 students (16-18yrs) [18] had significantly more contacts than other grades during closures, particularly late in the week. Because many Grade 12 students were not regularly attending classes at the school prior to the outbreak, they may have felt that they or their friends had not been exposed to the infection. One study [22] found that children aged twelve and over were significantly more likely to go to fast food restaurants and parties but less likely to go grocery shopping than children under twelve.

Conversely, one study [25] found that younger children were more likely to leave the home during a closure, with 53.2% of kindergarten pupils, 42.5% of primary school pupils, 30.3% of junior high school pupils and 33.2% of high-school pupils reporting that they left the house at least once. Primary school pupils were significantly more likely to leave the home to visit a supermarket or convenience store (likely with their parents), while junior high school pupils and primary school pupils were significantly more likely to leave the home to attend extracurricular studies compared with pupils in other school categories. It is unclear why this study shows the reverse of many others, but as this was the only study from Japan it may relate to cultural differences.

#### District

There is a small amount of evidence that location can affect the out-of-home activities children take part in during school closures, from a study of behaviour in children from two different school districts [27]. In this study, children in Jujuy were more likely to attend religious events, use public transport, and go to plazas and recreation areas than children in Ushuaia; children in Ushuaia were more likely to go to the movie theatre and restaurants than children in Jujuy. Socio-economic differences may well be the reason for this: Ushuaia has one of the lowest poverty rates in the country, whereas Jujuy has one of the highest.

#### Employment status of adults in the household

One study [24] found that if all adults in the home were employed, ill children were less likely to leave the home. A baseline household had a 34% probability of a child visiting any other venue; if all adults were employed, the probability of children leaving the home decreased to 24%. This is an unexpected finding in that we would expect that children living only with employed adults might have to leave the house for childcare arrangements; the authors did not offer reasons for the association between employed adults and reduced likelihood of children leaving the home.

#### Perceived appropriateness of school closure

Two studies found that parental opinion about the appropriateness of the school closure was significantly correlated with student participation in activities outside the home [19, 25]. Students of parents who thought the school closure was not appropriate reported a mean of 4.7 out-of-home activities, compared with a mean of 4.3 activities for students of parents who were unsure and 2.8 for students of parents who thought the closure was appropriate [19]. This pattern persisted when the analysis was restricted to the 202 students who were asymptomatic. Similarly, Mizumoto et al. [25] found that proportionately fewer children left the home in households that believed the closure was appropriate: 38.8%, compared to 53.2% of children in households who felt the closure was inappropriate.

#### Extent of closure

One study [25] found that extent of the school closure was significantly associated with the frequency of children leaving the home: closure of the entire school, closure of a single grade or single class suspension were associated with 47.8%, 32.2% and 40.3% of children leaving the home, respectively.

#### Length of time advised to isolate

One study, with high compliance rates with advised quarantine [28] found that children stayed at home for more than 94% of the days they were advised to be in quarantine and this figure was not associated with the length of quarantine nor did it fluctuate over the course of the quarantine period.

#### Day of the week

In one study [28] contact rates of uninfected students at the end of the week were lower than at the beginning. Contacts substantially increased for older children (grades 11 and 12) on Friday and Saturday.

#### Special childcare arrangements

Children in households where special childcare arrangements were needed during closure were more likely to leave the home than households in which children were independent and able to take care of themselves (53.1% versus 35.9%) [25].

#### Other factors considered

Child’s gender, household educational level, household income and household size were not associated with the likelihood of the child leaving the home during school closure [25].

### Parental attitudes towards school closure

#### Perceived benefit of closure

Parents generally agreed with school closures. Several studies reported high rates of parents being at least moderately supportive of the closure: 97% [29], 93% [25], 91% [22], 78% [27], 73% [30], and 71% [31]. The main reasons for agreeing with school closures were believing that: it would protect the health of the community, of the children themselves, and of the household; and believing that there were too many sick children for the school to remain open [19, 22, 25, 29]. Timperio et al. [29] found that over 90% of parents felt it was important to disinfect the schools while closed to reduce the community spread of influenza.

#### Perceived risk of infection

Several of the main reasons for disagreeing with school closures appeared to be related to perceived risk: parents cited beliefs that closures do not protect against influenza [27]; beliefs that the illness is only mild [19, 25]; and beliefs that school closure is not an effective measure against infection [25].

#### Practicalities of school closure

The other main reasons for disagreeing with school closures were related to the practicalities and subsequent impact of the closure: for example, concerns about the impact on the child’s education [27]; difficulties making childcare arrangements [25]; and concerns about the economic impact [19, 22]. Parents reported various difficulties associated with school closures, primarily lost income, the effort of arranging alternate childcare, and uncertainty about the duration of the closure [32, 33]. Some studies also illustrated a lack of consistency by schools regarding the importance of not participating in social activities: for example, 17% of parents reported that after-school activities were not cancelled [31] while others expressed concern that school athletic events were still held on days that school was closed [29].

## Discussion

During a major infectious disease outbreak, school closure has the potential to slow the spread of infection. The effects of a closure will be attenuated if children continue to mix. Of the 19 papers that we identified, all of them reported that some degree of mixing continued to occur outside of the home. We should not be surprised at this. Even for adults, self-isolation can be difficult [34] and stressful [35] and children often have wider social circles and feel more social pressure to interact. The precise extent to which contact patterns change during a closure is harder to determine. Only a limited number of studies have attempted to quantify this, reporting reductions in contacts from 70.3 on typical school days to 24.8 [16] and 14.2 to 6.5 [17]. The key variation in these number of absolute contacts are social and cultural differences but also technical definitions of what a contact is (such as physical contact or transitory contact).

Complicating matters is that the qualitative nature of contacts also changes. Many studies have explored what types of activities children engage in outside of the home during a closure. These include the full range of recreational and social activities, from shopping to meeting friends indoors, using public transport and visiting restaurants. It is likely that the type of activity is important in determining the likelihood of infection spreading. For example, participation in sports events have been noted to be particularly associated with the spread of influenza as have social events such as parties, whereas we can, perhaps, be less concerned about visits to a park [19]. Further research is needed on quantifying the rates of contact associated with the various different activities reported in this review; contacts in households, schools and workplaces are likely for a more sustained duration than in more transient social settings (such as shopping) but parties may form a middle ground. Assuming infection given a contact is a function of duration and type of contact, this can form the basis of more evidence-based modelling and risk assessment.

Reassuringly, our review found that relatively few children required special arrangements to be made for their childcare that might actively increase the risk of disease transmission, such as being placed into a semi-formal childcare arrangement with other children or being looked after by grandparents. The proportion of children left home alone unsupervised, however, while low, is of concern. If school closures are considered in the future, public health officials should consider how best to support parents and prevent this from occurring.

Parental attitudes associated with agreeing or disagreeing with school closures were similar to those seen in relation to other preventive health behaviours for infectious diseases [36, 37]. In particular, two of the studies included in this review suggested there was a strong association between allowing children to socialise outside the home and disagreeing with the school closure [19, 25]. Ensuring parents understand why school closure is important will be a key factor determining the success of the measure in any future disease outbreak. In this regard, it was concerning that two studies appeared to highlight a lack of clarity in terms of advice about children’s social activities and knowing what children were and were not advised to do [29, 31]. Advice from schools should be consistent with public health advice – hosting extra-curricular activities and sporting events during a closure sends mixed messages to parents and can be confusing or detrimental [14].

We found unclear evidence about the majority of the other predictors of out-of-home activities which we reviewed. In particular, there was mixed evidence about whether children showing symptoms of illness or who have been ill during the closure will take part in similar out-of-home activities to children who are not ill. Clearly, it would be particularly concerning if even symptomatic children are participating in out-of-home activities.

Different studies found that both older age and younger age were associated with leaving the house during school closures. It may be that the direction of findings depends on the activity in question. For example, younger children seem to be more likely to go grocery shopping (likely because they are too young to be left at home alone when their caregiver goes to the shops) whereas older children are more likely to take part in social activities like parties and going to restaurants.

In terms of how our findings fit with the wider literature, one particular discrepancy is worthy of note. Evidence from studies in which people are asked how they would react to a hypothetical school closure often find that parents believe they would co-operate with public health advice. For example, one study involving a hypothetical scenario of schools closing for three months due to an influenza pandemic found that 85% of parents responsible for children aged between 5-17 believed they would be able to keep their children from taking public transport, going to public events, and gathering outside the home during this lengthy school closure period [38]; another found that 96.7% of parents claimed they would keep their children away from others for a month if schools and child-care facilities were closed [39]. Despite these good intentions, our review of real school closures suggests parents are less likely to achieve this, even when schools are closed for much shorter periods of time. Regardless of the conviction with which people answer questions about their likely future actions, substantial caution is needed in using such data. The duration of planned closure of schools is perhaps critical here too; short closures of up to a couple of weeks may be manageable by parents as seen in the studies found but longer closures required for curtailing pandemic waves of the order of months may provide more challenge to them.

Further research is needed to identify how best to ensure that children are incentivised to stay at home during a school closure. The relatively sparse research conducted to date (limited by the real-world occurrence of school closures and the feasibility of conducting rapid research when these do occur) do not allow us to provide a ready answer to this question, but improved communication with both parents and children is likely to be required.

In terms of limitations for this review, the generalisability of the individual studies we identified is unclear. In particular a lot may depend on the cultural context, perceptions of the illness in question, the length of the closure, the socio-economic status of the families that are affected and the information or instructions that are given to them by public health authorities. With relatively few studies in this field, it is difficult to disentangle these effects.

Recent reviews of the incorporation of human behaviour into infectious disease models have advocated the use of appropriate, detailed, real-world behavioural data within infectious disease modelling [40, 41]. It is to be hoped that our identification of real-world data concerning social contact and mixing behaviour during unexpected school closures will help improve existing models and also promote more rigorous quantitative research in this area.

## Data Availability

None available. This paper is not based on original data.

---

**Supplementary Appendix 1.** Full details of activities outside the home and contacts with non-household members during school closures. This supplementary material is hosted by Eurosurveillance as supporting information alongside the article ‘The impact of unplanned school closure on children’s social contact: Rapid evidence review’, on behalf of the authors, who remain responsible for the accuracy and appropriateness of the content. The same standards for ethics, copyright, attributions and permissions as for the article apply. Supplements are not edited by *Eurosurveillance* and the journal is not responsible for the maintenance of any links or email addresses provided therein.

| **Study: Author (year), country** | **Participants** | **Activities outside the home and contacts with non-household members** |
| --- | --- | --- |
| Basurto-Davila et al. (2013), Argentina [27] | 226 households; Children from 3 schools closed for 2 weeks due to H1N1 (School A – children aged 6-12; School B – children aged 13-15; School C – children aged 6-15. Schools A and B were in the city of Ushuaia; School C was in Jujuy) | 67% of children visited public places at least once; 45% left the house several times. Approximate percentages taking part in out-of-home activities (Ushuaia region / Jujuy region respectively):<br>~45% / ~35% visited the supermarket;<br>~20% / ~40% visited plazas / recreation areas;<br>~28% / ~30% visited shopping malls;<br>~28% / 22% attended indoor friend gatherings;<br>~5% / ~30% attended religious events;<br>~10% / ~28% used public transport;<br>~20% / ~3% visited movie theaters;<br>~10% / ~12% attended indoor sports practice;<br>~15% / ~8% visited restaurants;<br>~3% / ~2% participated in group activities.<br>Childcare arrangements (Ushuaia region / Jujuy region respectively):<br>82% / 88% were left with a relative or family friend;<br>13% / 5% were with a hired nanny;<br>3% / 4% had ‘other special arrangements’;<br>2% / 1% of children were left alone. |
| Borse et al. (2011), USA [24] | 554 households; Median age of children: 8 years; Schools closed for 5-7 days due to | 30% of students visited at least one locale outside their homes.<br>For children without an influenza-like illness:<br>~20% visited a playground or park;<br>~5% went to a grocery store / mall / ran |
|  | H1N1 | errands;<br>~3% visited family;<br>~3% visited library,<br>~3% went to a restaurant;<br>~3% attended a religious function or service;<br>~2% attended a sports practice or game;<br>~2% went to a movie theatre; and friend's house.<br>~1% went to a friend's house;<br>~1% visited a doctor.<br>Children who were sick with an influenza-like illness before or during the closure were only slightly less likely to visit a playground or park; their second most common venue was a doctor visit which they were significantly more likely to visit than a non-sick child; and they were slightly more likely to visit family than non-sick children. |
| Braunack-Mayer et al. (2013), Australia [14] | 4 school principals, 25 staff, 14 parents, 13 students;<br>Students ranged in age from 12-17;<br>Schools either partially or fully closed due to H1N1 (length of closure unclear) | Qualitative study indicating most people adhered to advised quarantine, but in the absence of clear instructions, many invented their own rules.<br>Some parents quarantined their children to avoid being seen as irresponsible. However, many were home alone and so it was unclear whether children complied with this. Others reported seeing the closure as ineffective and did not quarantine their children. One student reported meeting friends regularly even though his parents believed he was at home. |
| Chen et al. (2011), Taiwan [30] | 232 households;<br>School for children aged 5-12;<br>School closed for 7 days due to H1N1 | 13% went to public places or gatherings at least once;<br>12% visited relatives;<br>5% went to parents' workplace.<br>60% were cared for by parents, 35% by other relatives, 4% by others, 1% stayed home alone. (Compared to pre-closure: 39% were cared for by parents after school, and 29% were in afterschool childcare.) |
| Effler et al. (2010), Australia [19] | 233 households;<br>Median age of | 74% participated in activities outside the home on at least one occasion, reporting a total of |
|  | children: 11 years (range 5-13);<br>3 schools closed due to H1N1;<br>School A closed entirely 'for the coming week';<br>Schools B and C cancelled classes only for grade 5 and grades 5-7 respectively | 860 out-of-home activities with an average of 3.7 out-of-home activities for each student. Activities reported (n during the week / n at weekend):<br>Park or beach (144 / 52);<br>Sporting event (86 / 128);<br>Grocery store (68 / 35);<br>Shopping mall (63 / 28);<br>Unspecified event or activity (58 / 17);<br>Party (28 / 37);<br>Music or art lesson (19 / 8);<br>Restaurant (13 / 14);<br>Cinema (16 / 5);<br>Religious service (1 / 16);<br>Sleepover (10 / 6);<br>Tutoring lesson (6 / 2).<br>19% of 202 asymptomatic students were cared for in a setting with children other than their siblings; 6% of 31 ill students were cared for with children other than their siblings. 10% cared for themselves at home for at least a portion of the closure period. |
| Epson et al. (2015), USA [20] | 35 households, representing 67 students;<br>1 elementary school and 1 junior and senior high school that are housed in the same building complex;<br>Schools closed between 29.01.13 and 05.02.13 due to influenza-like illness | 58% visited at least one outside venue:<br>58% went to grocery stores;<br>33% went to restaurants;<br>25% went to sports practices;<br>25% went to religious services;<br>25% visited friends' houses.<br>Childcare involving contact with non-household members:<br>9% were looked after by an adult who lives outside the household;<br>6% went to work with their parents;<br>3% were in a childcare program.<br>9% were left home alone. |
| Gift et al. (2010), USA [23] | 214 households, representing 269 children under 18;<br>Elementary school closed for 1 week due to H1N1 | 69% visited at least one other location.<br>The most common activities outside of the home were: 'other'; shopping, sports activities, restaurants, visiting family, visiting healthcare provider, visiting friends, going to a movie or concert, visiting the library, |
|  |  | attending a religious venue, going to a party, and going to a drive-through restaurant.<br>Home was the primary location for 77%; the next most common location was another family member's home, followed by non-family member's home, parents' workplace, vacation, daycare, and 'other'. |
| Jackson et al. (2011), UK [16] | 107 students (only 46 reported how many times they visited public places during closures);<br>Children aged 11-15;<br>School closed for 1 week, reopened for 2 days, then closed for another week, due to H1N1 | 98% visited more than one place.<br>78% visited shops (compared to 94% when school was open);<br>59% visited a park or playing field (81% when school was open);<br>46% visited places of worship (66% when school was open);<br>43% used public transport (58% when school was open);<br>37% visited cafes (50% when school was open);<br>28% visited leisure centres (42% when school was open);<br>26% went to parties (22% when school was open);<br>17% visited movie theatres (17% when school was open).<br>Fewer students visited shops, places of worship, parks and playing fields at least once per week when school was closed than when open; for other places frequency of visits did not differ.<br>Among caregivers for whom information was available, 125/182 (69%) would have seen the student on a typical school day.<br>73 students provided their typical number of contacts per day during closure and 35 also provided information for a typical school day. Mean totals of reported contacts were 70.3 and 24.8 during typical school days and closure respectively. |
| Johnson et al. | 220 households, | 89% visited at least one public location despite |
| (2008), USA [22] | representing 355 children;<br>Median age of children: 12 (range 5-19);<br>Schools closed between 01.11.06 and 12.11.06 due to influenza virus B | recommendations to avoid large gatherings and 47% travelled outside of the county.<br>44% visited grocery stores;<br>35% visited fast food restaurants;<br>34% visited church services;<br>19% visited malls;<br>15% visited parties or sleepovers.<br>10% had to make special childcare arrangements including grandparents or other relatives or other adults, taking the child to work, having older siblings watch them or using childcare programs.<br>3% had to have their child spend more than one night outside the household for childcare purposes. |
| Litvinova et al. (2019), Russia [17] | 450 participants including students and their household members;<br>2 schools – School A for children aged 6-17 and School B for children aged 6-15;<br>Gradual school closure policies performed every year to mitigate spread of seasonal influenza;<br>Schools closed for 7 days | When school closure policy was in place, there was a significant reduction in the number of contacts made by students (14.2 contacts per day when open vs. 6.5 when school was closed). School-closure policies induced students to significantly reduce their number of contacts with individuals aged under 18 (75% reduction) and 19-59 year-olds (20% reduction), while simultaneously increasing contacts with individuals aged 60 or over (52% increase), although the absolute value remained low (i.e., less than one contact per day). |
| McVernon et al. (2011), Australia [28] | 314 households;<br>33 schools with children under 18;<br>Schools with confirmed cases of H1N1 in multiple classes were entirely closed for 7 days while schools with confirmed cases in | 43 households reported that a child spent at least one day outside the family home, and mixing with other children occurred on almost half of these occasions (48.8%), whether or not there was an influenza case in the family. Contact with children who were not immediate family members was far less likely during days spent at home. No child visited a household in which another child was ill, compared with reported child visitors in |
|  | only one class were instructed to close only that class | 15.9% of 226 homes without a case.<br>Compared to children in households that complied with recommendations to stay at home, children in households that did not comply with the recommendations were more likely to have been cared for during the quarantine period by an adult from outside the home (28.3% compared with 4.0% for compliant households; $p < 0.001$), thus also contravening the quarantine recommendation not to mix with adults from outside the household. This distinction was especially marked for households in which there was a confirmed case of influenza, where the difference was 44.4% of children receiving outside care in non-compliant households compared with 2.4% of those that were compliant. |
| Miller et al. (2010), USA [18] | 63 parents of 176 lower school students (grades 5-8);<br>188 upper school students (grades 9-12);<br>Week-long closure due to H1N1 | Mean number of days spent on activities (upper school):<br>3.42 any other outdoor activity;<br>2.44 eating at restaurants;<br>1.89 using public transport;<br>1.48 hosting a friend;<br>1.47 shopping;<br>1.47 any other indoor activity;<br>0.44 working at a job.<br>Average number of friends seen per day (upper school): 2.53 Wednesday, 2.06 Thursday, 2.59 Friday, 2.40 Saturday, 1.23 Sunday, 1.02 Monday, 1.05 Tuesday.<br>Proportion of caregivers (upper school): 0.62 parent, 0.24 sibling, 0.07 grandparent, 0.07 other, 0.06 nanny or babysitter, 0.07 friend's caretaker, 0.11 other, 0.88 self.<br>Mean number of days spent on activities (lower school):<br>2.77 any other outdoor activity;<br>1.34 eating at restaurants; |
|  |  | 1.12 any other indoor activity;<br>1.05 shopping;<br>0.73 visiting a friend;<br>0.55 hosting a friend;<br>0.10 using public transport.<br>Average number of friends seen per day<br>(lower school): 0.30 Wednesday, 0.52<br>Thursday, 0.84 Friday, 0.83 Saturday, 1.17<br>Sunday, 0.74 Monday, 0.68 Tuesday.<br>Proportion of caregivers (lower school):<br>0.85 parent, 0.30 sibling, 0.09 grandparent,<br>0.15 other family, 0.27 nanny or babysitter,<br>0.03 friend's caretaker, 0.06 other, 0.76 self. |
| Mizumoto et al.<br>(2013), Japan [25] | 882 households;<br>25.2% in<br>kindergarten, 24.8%<br>in primary school,<br>25.1% in junior<br>high school and<br>24.9% in high<br>school, age range 4-18;<br>'School closure or<br>class suspension at<br>least once' due to<br>H1N1 | 20.5% left the home for non-essential reasons.<br>Of the 351 students who left the home:<br>40.9% went shopping at a supermarket or<br>convenience store;<br>19.3% went shopping at a department store or<br>large shopping centre;<br>18.7% attended extra classes, prep school, or<br>English school;<br>9.5% visited an outdoor playground or pool;<br>8.7% participated in a sports activity;<br>8.2% ate at restaurants;<br>2.0% participated in a concert, drawing, or<br>other artistic activities;<br>1.0% had an overnight stay away from home;<br>0.5% participated in religious activities;<br>0.3% participated in an outdoor group activity;<br>0.3% visited the cinema;<br>0.1% went to a party;<br>11.2% took part in other activities outside the<br>house.<br>28.5% children looked after themselves;<br>64.3% were looked after by another household<br>member; 7.3% required a special arrangement<br>such as parental absence from work. |
| Russell et al.<br>(2016), USA [26] | 99 households,<br>representing 197<br>children; | 77% of children went outside the home or<br>visited with a non-household member,<br>participating in a mean of two activities (IQR |
|  | Students in pre-kindergarten up to 12<sup>th</sup> grade;<br>School closed for 4 days due to influenza-like illness | 1-4):<br>54% visited a mall or department store;<br>51% visited a grocery store;<br>32% attended religious services;<br>32% visited family;<br>24% went to restaurants or sports activities.<br>Childcare involving contact with non-household members:<br>20% were looked after by a non-household adult;<br>1% were in a childcare program. |
| Steelfisher et al. (2010), USA [31] | 523 parents;<br>Ages not reported;<br>Childcare centres and schools closed due to H1N1 - 10% were closed for 1 day, 19% for 2 days, 29% for 3 days, 15% for 4 days, 17% for 5 days, 9% for more than 5, 2% didn't know | 56% reported their child participated in at least one activity involving people outside the household:<br>30% visited other children's houses;<br>30% went grocery shopping;<br>23% went to fast food restaurants;<br>17% went to public events such as movies, sporting events or concerts;<br>15% visited large shopping areas or malls;<br>6% attended social events such as parties or dances.<br>81% were cared for by an adult in the household, 20% by a family member outside the household, 1% by a friend/neighbour, 3% by a professional care provider, and 10% stayed home alone. |
| Timperio et al. (2009), USA [29] | 262 households representing 480 children;<br>Ages not reported;<br>Two schools closed due to seasonal influenza; one closed for 3 days and the other closed for 4 days | 43.3% visited strip malls or Wal-Mart (the largest store in the area);<br>42.9% visited family;<br>38.7% went grocery shopping;<br>32.6% ate at restaurants;<br>30.3% either visited friends' houses or had friends visiting their house;<br>29.1% attended religious services;<br>23.8% took part in sports activities;<br>17.6% went to public gatherings such as concerts, movies or festivals;<br>8.4% went to a part time job. |
| Tsai et al. (2017), USA [32] | 208 households representing 423 | 3% of children were put into a childcare program; |
|  | students;<br>Children aged under 18;<br>School closed for 8 days due to influenza | 1% went to work with their parents.<br>1% were left at home without supervision and another 15% were old enough to care for themselves. |
| van Gemert et al. (2018), Australia [21] | 99 students with laboratory confirmed pH1N1; Age 6-17; 49% were aged 14-15; 33% were aged 16-17; 9% were aged 12-13; 5% were aged 10-11; 4% were aged 6-7; Seven schools closed for a range of 3-9 days (not including weekends) due to H1N1 | 26% (21/81) who reported usually taking part in extra-curricular activities (not sports or religious activities) continued to take part in extra-curricular activities;<br>23% (14/62) continued to participate in sports activities;<br>20% (2/10) continued to take university classes;<br>11% (2/18) continued to attend part-time work;<br>5% (1/20) continued to take part in religious activities. |
| Zheteyeva et al. (2017), USA [33] | 2,229 households representing 4,247 students;<br>Kindergarten – Grade 12;<br>Schools closed for 4 days in preparation for Hurricane Isaac | 5.3% of children went to work with their parents; 2.6% of children were put in a childcare program. 11.6% were old enough to care for themselves, while 2.5% were left at home without supervision. |

## Notes

**Funding statement** The research was funded by the National Institute for Health Research Health Protection Research Unit (NIHR HPRU) in Emergency Preparedness and Response at King’s College London in partnership with Public Health England (PHE), in collaboration with the University of East Anglia and Newcastle University. The views expressed are those of the author(s) and not necessarily those of the NHS, the NIHR, the Department of Health and Social Care or Public Health England. Dale Weston’s time on this project was funded by the National Institute for Health Research Health Protection Research Unit (NIHR HPRU) in Modelling Methodology at Imperial College London in partnership with Public Health England (PHE). IH is also supported by the NIHR Health Protection Research Unit in Modelling Methodology and the NIHR policy research programme Operational Research for Emergency Response and strategic planning Analysis (OPERA).

**Conflict of interest** None declared.

### Competing Interest Statement

The authors have declared no competing interest.

### Funding Statement

The research was funded by the National Institute for Health Research Health Protection Research Unit (NIHR HPRU) in Emergency Preparedness and Response at King’s College London in partnership with Public Health England (PHE), in collaboration with the University of East Anglia and Newcastle University. The views expressed are those of the author(s) and not necessarily those of the NHS, the NIHR, the Department of Health and Social Care or Public Health England. Dale Weston’s time on this project was funded by the National Institute for Health Research Health Protection Research Unit (NIHR HPRU) in Modelling Methodology at Imperial College London in partnership with Public Health England (PHE). IH is also supported by the NIHR Health Protection Research Unit in Modelling Methodology and the NIHR policy research programme Operational Research for Emergency Response and strategic planning Analysis (OPERA).

